# Oncology Researchers’ and Clinicians’ Perceptions of Complementary, Alternative, and Integrative Medicine: An International, Cross-Sectional Survey

**DOI:** 10.1101/2024.03.03.24303667

**Authors:** Jeremy Y. Ng, Jassimar Kochhar, Holger Cramer

## Abstract

**Background:** Complementary, alternative, and integrative medicine (CAIM) has become an increasingly popular supportive therapy option for patients with cancer. The objective of this study was to investigate how researchers and clinicians in the oncology field perceive CAIM.

**Methods:** We conducted an online, anonymous, cross-sectional survey for researchers and clinicians who have published their work in oncology journals that are indexed in MEDLINE. The link to the survey was sent to 47, 991 researchers and clinicians whose contact information was extracted from their publications. The survey included various multiple-choice questions, and one open-ended question at the end to allow for any additional comments.

**Results:** 751 respondents completed the survey, and they mostly identified themselves as researchers (n=329, 45.13%), or as both researchers and clinicians (n=332, 45.45%) in the field of oncology. Over half of the respondents perceive mind-body therapies (n=354, 54.97%) to be the most promising CAIM category with regards to the prevention, treatment, and/or management of diseases related to oncology, and many respondents agreed that most CAIM therapies are safe (n=218, 37.39%), and that clinicians should receive training on CAIM therapies via formal (n=225, 38.59%) and supplemental education (n=290, 49.83%). However, many respondents were unsure when asked if most CAIM therapies are effective (n=202, 34.77%).

**Conclusions:** The findings from this study demonstrated great current interest in the use of CAIM in oncology. This information can serve as a foundation for conducting additional research and creating customized educational materials for researchers and clinicians in oncology.

## Background

Cancer, although commonly interpreted as a single illness, comprises over 100 different diseases, all of which develop from abnormalities within fundamental aspects of the cell growth cycle [1–5]. The global burden of cancer continues to increase, largely fuelled by aging populations and a greater prevalence of cancer-promoting behaviours, such as smoking [6]. The International Agency for Research on Cancer provides an evaluation of the global incidence and mortality of cancer through the GLOBOCAN estimate, which explores new cancer cases and cancer deaths for 36 cancer types throughout 185 countries or territories [7]. In 2020 alone, the GLOBOCAN estimate appraised the global prevalence of cancer as 19.3 million new cases and almost 10.0 million deaths [7]. Chemotherapy, radiotherapy, and surgery are among the most common types of cancer treatments, and novel approaches to cancer management such as immunotherapy, stem cell therapy, and nanoparticles are continuously advancing [8–9]. Although essential for curative cancer treatment, these treatment strategies pose a vast array of adverse effects, are both physically and emotionally taxing on patients [9]. Therefore, due to negative adverse effects associated with existing cancer therapies, patients with cancer may be increasingly inclined to explore supportive management strategies, such as complementary, alternative, and integrative medicine (CAIM).

CAIM usage has experienced increasing popularity among patients in recent years, and individuals battling cancer bode no exception [10–14]. The US National Center for Complementary and Integrative Health (NCCIH) has defined “complementary medicine” as a non-mainstream approach used *together with* conventional medicine, whereas “alternative medicine” is defined as a non-mainstream approach used *in place* of conventional medicine [15]. Moreover, “integrative medicine” is the coordination of conventional and complementary approaches, thus emphasizing the use of multimodal interventions to develop combinations of health approaches centred around holistic care/treatment [15–17]. For the purpose of this study, we will collectively refer to this group of diverse therapies as CAIM. Examples of CAIM include acupuncture, mind-body practices such as yoga, herbal supplements, music/art therapy, and chiropractic therapy [15–22]. There has been an increase in CAIM usage by patients with cancer in order to moderate side effects of invasive treatments (chemotherapy and radiation) and for perceived benefits such as health promotion, disease symptom management, illness prevention, and immune function improvement [23–25]. For example, a 2011 meta-analysis conducted by Horneber et al. reported that 49% of patients with cancer throughout 18 selected countries currently used CAIM [26], and many studies have found that patients with cancer use it as a supportive measure to alleviate associated symptoms or the side effects of curative therapy [27–34].

However, despite the relatively high prevalence of CAIM use among patients with cancer, healthcare professionals lack knowledge about the safety and efficacy of CAIMs, and current literature lacks evidence-based information regarding the safety, efficacy, and benefits of CAIMs in oncological settings. As a result, healthcare professionals are often unequipped to discuss the potential harms and benefits of CAIMs with patients and lack guidance pertaining to CAIM recommendations in oncology clinical practice guidelines [35]. This has led to inconsistent guidance from physicians regarding the integration of CAIMs into conventional healthcare settings [36]. This has also led over half of patients with cancer to avoid discussing their use of CAIM with clinicians due to concerns such as clinicians’ lack of knowledge and interest regarding CAIM [37–41].

Therefore, given the growing prevalence of CAIM among patients with cancer and the debates surrounding its validity within healthcare, it is evident that a better understanding of the perceptions of oncology medical researchers and clinicians towards CAIM is required. An exploration of this sort would highlight the reservations and support that oncology clinicians and researchers demonstrate towards CAIM and may dismantle the gaps in current literature pertaining to clinicians’ and researchers’ perspectives on CAIM. Therefore, the objective of this study is to collect the perceptions of oncology researchers and clinicians regarding CAIM.

## Methods

### Transparency Statement

Approval was granted by the Research Ethics Board at the University Hospital Tübingen before beginning this project (REB Number: 389/2023BO2). Prior to recruiting participants, the study protocol was registered and made available on the Open Science Framework (OSF) [42].

### Study Design

The study was carried out as an online, anonymous, cross-sectional survey targeting researchers and clinicians who have published in oncology medical journals indexed in MEDLINE [43].

### Sampling Framework

A complete sample of corresponding authors who have published articles in oncological journals indexed in MEDLINE within approximately the past 3 years (between August 1^st^, 2020, and May 1^st^, 2023) were chosen. The selection involved all cancer journals [44] indexed on https://journal-reports.nlm.nih.gov/broad-subjects/. The NLM IDs of the selected journals were first extracted, after which a search strategy based on such IDs was completed on OVID Medline. The resulting list of PubMed IDs (PMIDs) identified through the search was exported as a .csv file in batches of 2000 records at a time and inputted into an R script in batches of 200 PMIDs at a time. The R script subsequently retrieved authors’ names, affiliation institutions, and email addresses (developed based on the easyPubMed package [45]). The study included authors who had published manuscripts of any type.

### Participant Recruitment

The sampling framework was used to generate a contact list comprising of individuals primarily assumed to be oncology researchers and clinicians based on their publication of articles in oncology journals indexed in MEDLINE. It was expected that the curated list contained duplicate email addresses due to participants potentially contributing/authoring multiple manuscripts within our sample, thus, duplicate addresses were removed from the dataset prior to recruitment emails being sent out. Furthermore, to account for any incorrect author names retrieved during our search, we attempted to correct such (where possible) through a Google search, rather than completely omitting the incorrectly retrieved name from our sample. The platform that was used to both send the emails and create the survey itself was SurveyMonkey [46]. An email which outlined the study’s objectives and provided a survey link was sent to potential participants. Upon clicking the survey link, participants were directed to the initial page of the survey. Participants were prompted to confirm their consent to the specified terms and conditions linked to survey participation. Only those who responded with a “Yes” were granted access to view and respond to the survey questions. Participants were sent reminder emails during the first, second, and third weeks following the initial invitation email, after which participants were provided with a total of four weeks after the third (final) reminder to complete the survey. The survey was open from August 29, 2023 to November 13, 2023. Participants were able to skip any questions within the survey that they do not wish to answer.

### Survey Design

The first page of the survey asked participants a screening question, which was followed by a series of demographic questions. Throughout the rest of the survey, participants were asked to provide their perceptions on CAIM and a multiple-choice format was used for the majority of the survey questions. Two independent CAIM researchers pilot tested the survey by providing their feedback to the questions detailed within such prior to its distribution. A copy of the survey can be found on the following link: https://osf.io/kb2zp.

### Data Management and Analysis

Quantitative data obtained from the multiple-choice questions were analyzed and used to generate basic descriptive statistics, such as counts and percentages. Furthermore, the qualitative data from the one open-ended question were analyzed through a thematic content analysis [47]. This entailed the responses being interpreted and being assigned a distinct code, which was a representation of the main component of their response.

## Results

### Demographics

In total, 47 991 emails were sent, with 25 651 being unopened while 6605 bounced. The survey had 751 responses in total (1.8% response rate of unopened and opened, and 4.7% of just the opened). Raw survey data is available at https://osf.io/a94rp. The survey took approximately 9 minutes to complete. Approximately the same number of respondents indicated that they self-identify as a researcher (n=329, 45.13%), and as both a researcher and a clinician (n=332, 45.45%) in the field of oncology. In terms of World Health Organization World Regions, the majority of respondents were from Europe (n=315, 45.32%) and the Americas (n=210, 30.22%). Over half of respondents described themselves as faculty members/principal investigators (n=356, 51.22%), with the next most common being clinicians such as physicians, nurses, etc (n=288, 41.44%). Most respondents also described themselves as a senior researcher or clinician with >10 years of starting their career post formal education (n=417, 60%). The primary research area of most respondents was clinical research (n=416, 68.53%). The full information regarding participant characteristics can be found in **Table 1**.

**Table 1:** Characteristics of Survey Participants.

|  |  |
| --- | --- |
| **Sex (n=695)** |  |
| Male | 335 (48.20%) |
| Female | 346 (49.78%) |
| Intersex | 0 (0.00%) |
| Prefer not to say | 14 (2.01%) |
| Prefer to self describe | 0 (0.00%) |
| **Age (n=696)** |  |
| Under 18 | 0 (0.00%) |
| 18-24 | 1 (0.14%) |
| 25-34 | 91 (13.04%) |
| 35-44 | 206 (29.60%) |
| 45-54 | 189 (27.16%) |
| 55-64 | 138 (19.83%) |
| 65 or older | 60 (8.62%) |
| Prefer not to say | 11 (1.58%) |
| **Visible Minority (n=691)** |  |
| Yes | 112 (16.21%) |
| No | 545 (78.87%) |
| Prefer not to say | 34 (4.92%) |
| **World Health Organization World Region (n=695)** |  |
| Africa | 10 (1.44%) |
| Americas | 210 (30.22%) |
| Eastern Mediterranean | 22 (3.17%) |
| Europe | 315 (45.32%) |
| South-East Asia | 74 (10.65%) |
| Western Pacific | 46 (6.62%) |
| Prefer not to say | 18 (2.59%) |
| **Current Position (n=695)** |  |
| Clinician Student | 1 (0.14%) |
| Clinician | 288 (41.44%) |
| Graduate student | 27 (3.68%) |
| Postdoctoral fellow | 50 (7.19%) |
| Faculty member/principal investigator | 356 (51.22%) |
| Research support staff | 20 (2.88%) |
| Scientist in academia | 164 (23.60%) |
| Scientist in industry | 9 (1.29%) |
| Scientist in third sector | 12 (1.73%) |
| Government scientist | 14 (2.01%) |
| Other | 18 (2.59%) |
| **Career Stage (n=695)** |  |
| Graduate or clinician student | 11 (1.58%) |
| Early career researcher or clinician (<5 years post education) | 118 (16.98%) |
| Mid-career researcher or clinician (5-10 years post education) | 149 (21.44%) |
| Senior researcher or clinician (>10 years post education) | 417 (60.00%) |
| **Primary research area (n=607)** |  |
| Clinical research | 416 (68.53%) |
| Preclinical research – in vivo | 121 (19.93%) |
| Preclinical research – in vitro | 116 (19.11%) |
| Health systems research | 57 (9.39%) |
| Health services research | 120 (19.77%) |
| Methods research | 70 (11.53%) |
| Epidemiological research | 117 (19.28%) |
| Other | 41 (6.75%) |

### Complementary, Alternative, and Integrative Medicine

Most respondents had never conducted research in any area of CAIM (n=415, 68.82%). Over half of the respondents perceived mind-body therapies (n=354, 54.97%) to be the most promising CAIM category with regards to the prevention, treatment, and/or management of diseases related to oncology, with biologically based practices being the next most promising (n=278, 43.17%) (**Figure 1**). Many respondents also declared that their patients have sought counselling or disclosed using biologically based practices (n=270, 79.18%) and whole medical systems (n=224, 65.69%). It was also indicated by respondents that most commonly, only 0-10% (n=112, 33.04%) of their patients disclosed that they used CAIM or asked for their counselling on CAIM. When asked in which area of CAIM the respondents have practiced or recommended to their patients, most said mind-body therapies (n=139, 40.76%), and that they have not practiced or recommended CAIM to their patients (n=127, 37.24%). A high number of participants also said that they had not received any formal (n=269, 79.35%) or supplemental (n=223, 65.98%) training in any of the areas of CAIM. The majority of respondents indicated that the resource they used to learn more about CAIM was academic literature (n=580, 89.78%). When asked about CAIM in general and to what degree they agreed with a series of statements, some respondents agreed that most CAIM therapies are safe (n=218, 37.39%), that there is value to conducting research on CAIM therapies (n=307, 52.66%), that more research funding should be allocated to study CAIM therapies (n=224, 38.42%), and that clinicians should receive training on CAIM therapies via formal (n=225, 38.59%) and supplemental education (n=290, 49.83%) (**Figure 2**). However, respondents neither agreed nor disagreed when asked if most CAIM therapies are effective (n=202, 34.77%), if most CAIM therapies should be integrated into mainstream medical practice (n=164, 28.13%), and if insurance companies should cover the cost of most CAIM therapies (n=198, 34.08%). Respondents disagreed when asked if they would be comfortable counselling their patients about most CAIM therapies (n=96, 31.27%), and if they would be comfortable recommending most CAIM therapies to their patients (n=101, 33.01%).

**Figure 1:**
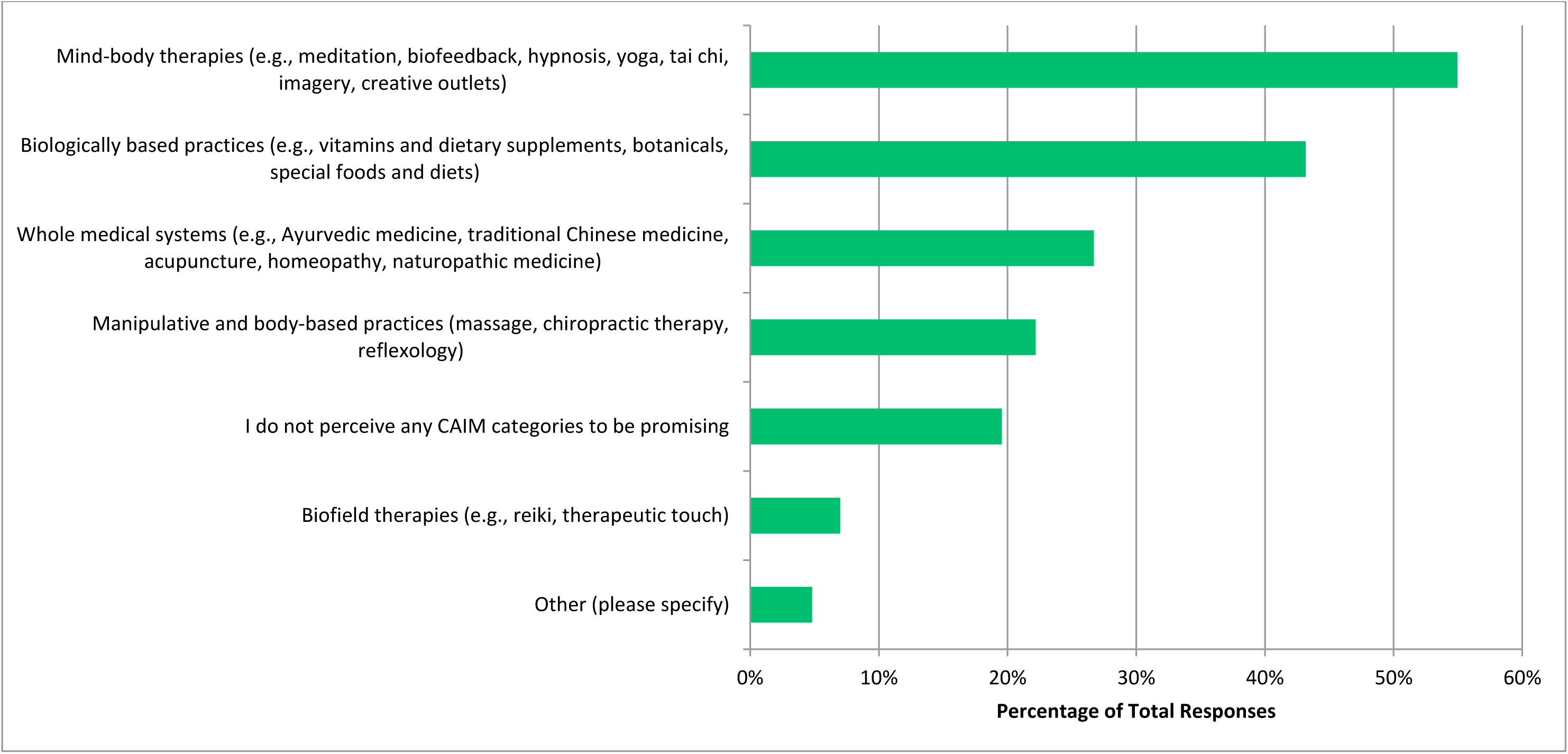
CAIM Category Perceived to be the Most Promising in Oncology.

**Figure 2:**
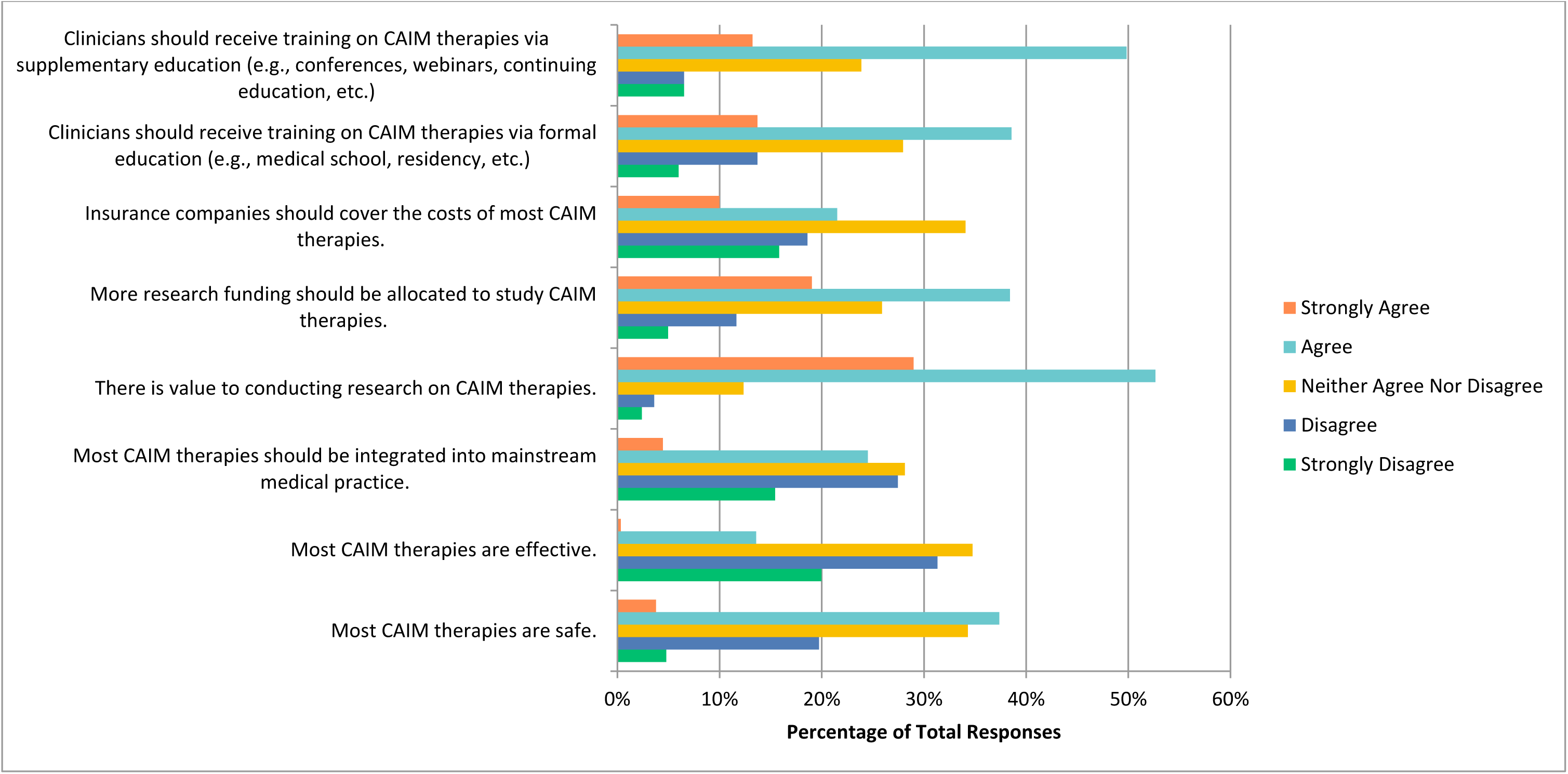
Agreement with the Following Statements Regarding CAIM in General.

### Mind Body Therapies

When asked about mind-body therapies such as meditation, biofeedback, hypnosis, and yoga, most respondents agreed that this therapy is safe (n=329, 56.63%), that there is value to conducting research on this therapy (n=323, 55.79%), that more research funding should be allocated to study this therapy (n=242, 41.72%), that clinicians should receive training on this therapy via formal (n=231, 39.83%) and supplemental education (n=299, 51.64%).

Conversely, respondents were unsure, and neither agreed nor disagreed that mind-body therapies are effective (n=266, 45.86%), that most mind-body therapies should be integrated into mainstream medical practice (n=208, 35.80%), and that insurance companies should cover the cost of mind-body therapies (n=216, 37.31%) (**Figure 3**). Some respondents also agreed that they would be comfortable counselling their patients about most mind-body therapies (n=95, 31.15%), and that they would be comfortable recommending most mind-body therapies to their patients (n=90, 29.70%).

**Figure 3:**
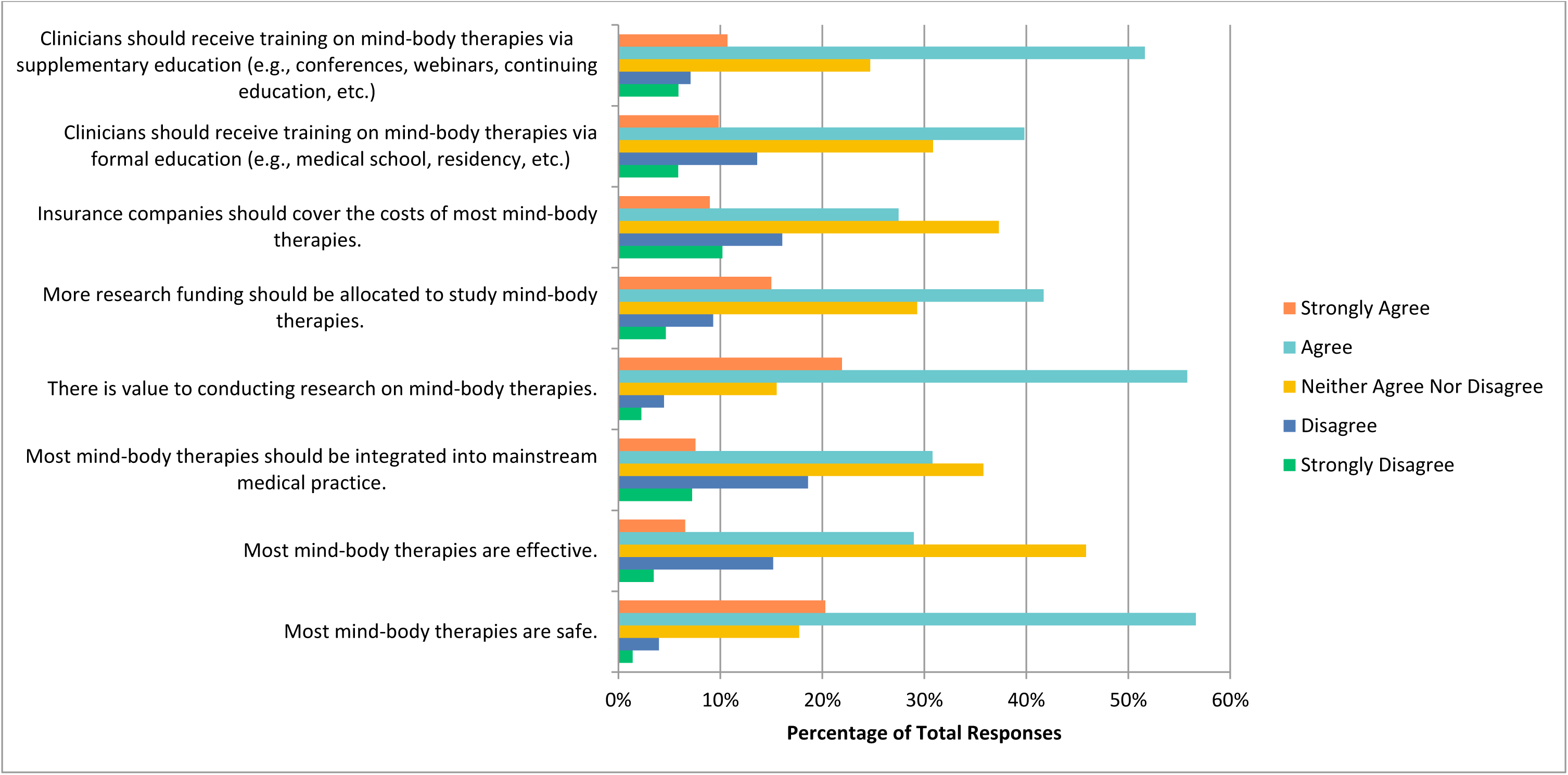
Agreement with the Following Statements Regarding Mind-Body Therapies.

### Biologically Based Practices

Next, participants were asked about biologically based practices such as vitamins and dietary supplements, botanicals, and special foods. Respondents were in agreement that there is value to conducting research on this therapy (n=313, 54.34%), that more research funding should be allocated to study this therapy (n=223, 38.99%), that clinicians should receive training on this therapy via formal (n=211, 36.70%) and supplemental education (n=239, 41.71%).

Respondents were undecided, and neither agreed nor disagreed that most biologically based practices are safe (n=217, 37.67%), effective (n=226, 39.44%), should be integrated into mainstream medical practice (n=193, 33.51%%), and that insurance companies should cover the cost of most biologically based practices (n=224, 38.96%) (**Figure 4**). They also disagreed when asked if they would be comfortable counselling (n=80, 26.32%) and recommending (n=96, 29.18%) most biologically based practices to patients.

**Figure 4:**
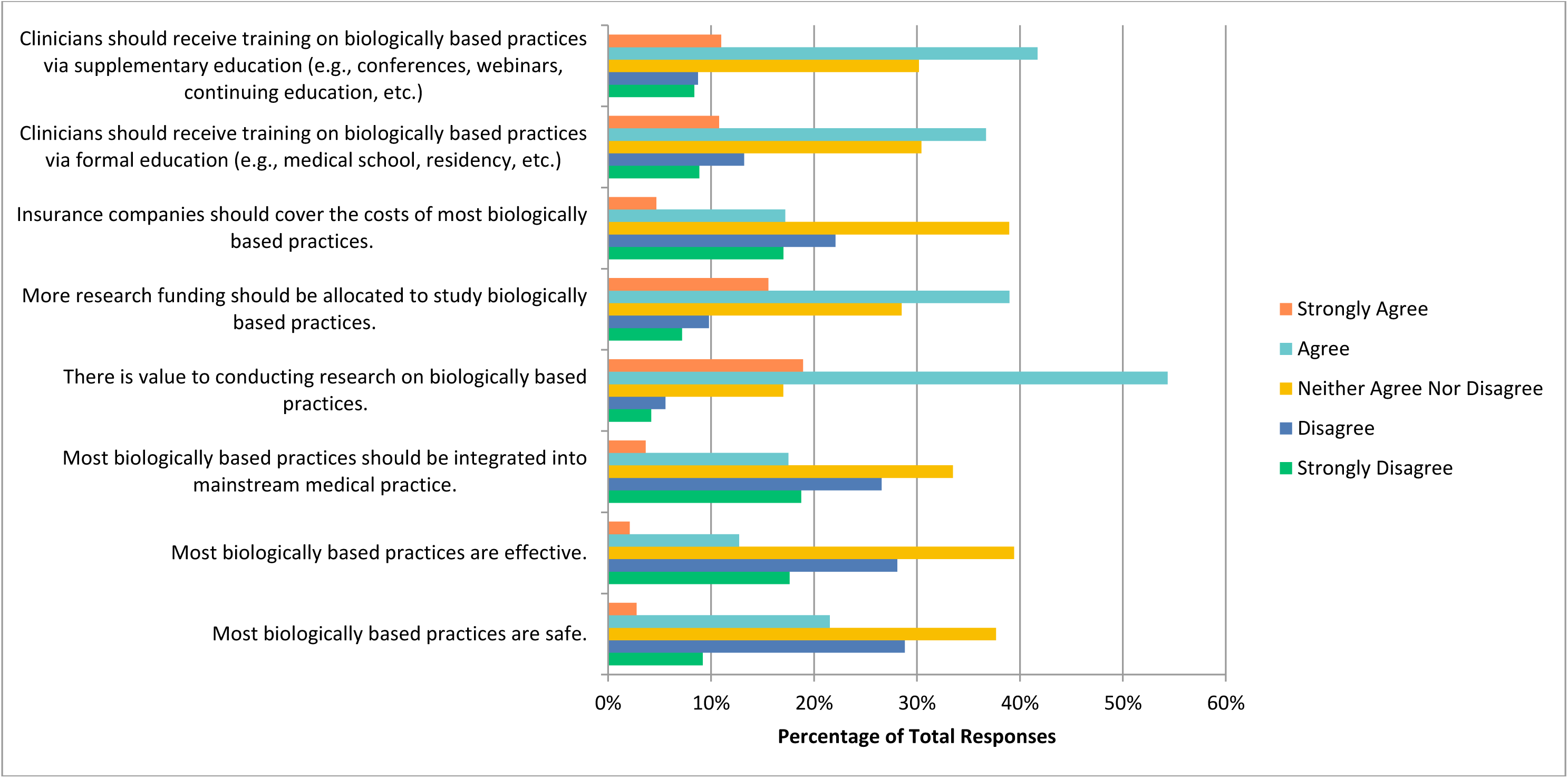
Agreement with the Following Statements Regarding Biologically Based Practices.

### Manipulative and Body-Based Practices

The next CAIM therapy that participants were asked about was manipulative and body-based practices, which encompasses massage, chiropractic therapy, and reflexology. Participants agreed that there is value to conducting research on this therapy (n=272, 47.47%) and that clinicians should receive training on this therapy via supplementary education (n=224, 39.23%). However, they were uncertain, and neither agreed nor disagreed when asked if most manipulative and body-based practices are safe (n=218, 37.98%), effective (n=238, 41. 54%), that they should be integrated into mainstream medicine (n=226, 33.37%), that more research funding should be allocated to study this therapy (n=196, 34.27%), that insurance companies should cover the cost of this therapy (n=215, 37.52%), and that clinicians should receive training via formal education (n=191, 33.39%) (**Figure 5**). Respondents were also unsure and neither agreed nor disagreed about if they would be comfortable counselling patients about most manipulative and body-based practices (n=95, 31. 35%), and disagreed that they would be comfortable recommending most manipulative and body-based practices to their patients (n=94, 30.92%).

**Figure 5:**
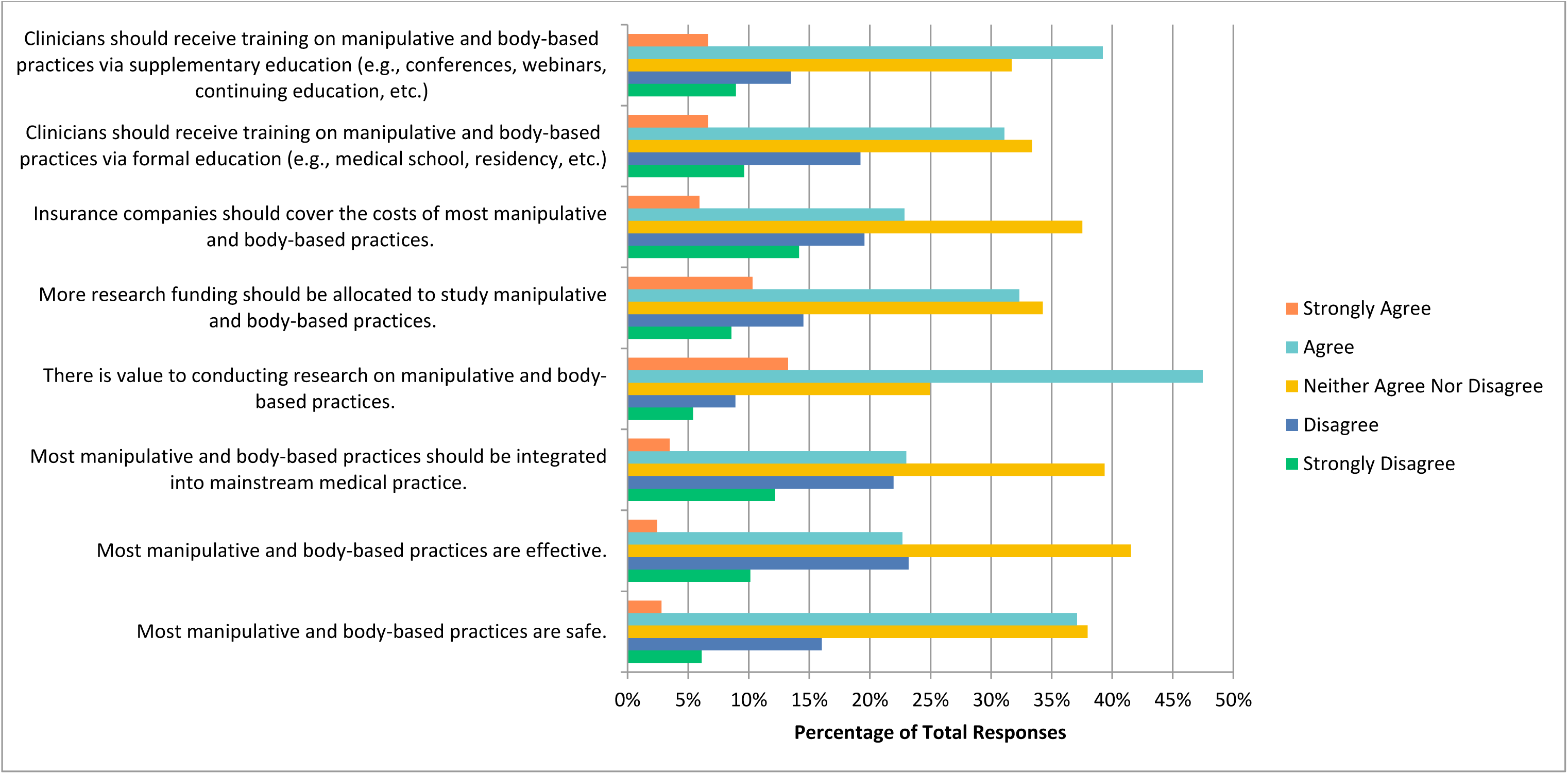
Agreement with the Following Statements Regarding Manipulative and Body-Based Practices.

### Biofield Therapies

Biofield therapies include practices such as Reiki and therapeutic touch. Respondents only agreed to one statement for this therapy, which is that there is value to conducting research on this topic (n=200, 35.03%). However, they were divided and neither agreed nor disagreed when asked if they believe most biofield therapies are safe (n=262, 45.96%), effective (n=259, 45.52%), that they should be integrated into mainstream medical practice (n=225, 39.40%), that more research funding should be allocated to study this therapy (n=192, 33.68%), that insurance companies should cover the cost of these therapies (n=226, 39.65%), and that clinicians should receive training on this therapy via formal (n=199, 34.97%), or supplemental (n=212, 37.32%) education (**Figure 6**). Respondents also disagreed that they would be comfortable counselling patients about most biofield therapies (n=98, 32.45%), and strongly disagreed (n= 96, 32.00%) and disagreed (n=96, 32.00%) in equal percentages when asked if they would be comfortable recommending biofield therapies to patients.

**Figure 6:**
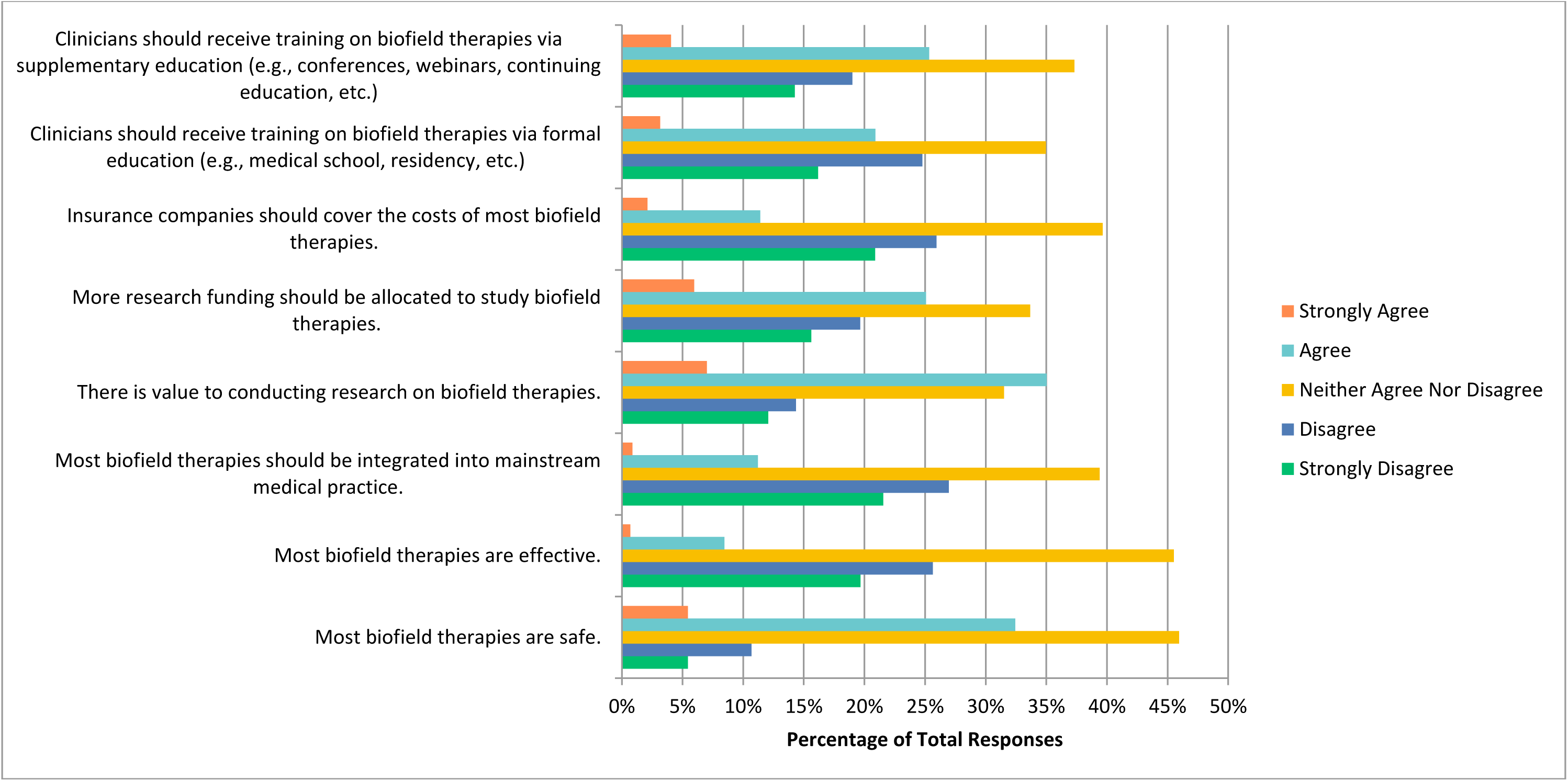
Agreement with the Following Statements Regarding Biofield Therapies.

### Whole Medical Systems

Respondents were finally asked about whole medical systems, which includes Ayurvedic medicine, traditional Chinese medicine, acupuncture, homeopathy, and naturopathic medicine. The respondents agreed that there is value to conducting research on this therapy (n=268, 46. 94%), that more research funding should be allocated to study this therapy (n=195, 34.21%), and that clinicians should receive training on this therapy via supplemental education (n=210, 37.04%). They were however unsure and neither agreed nor disagreed when asked if most manipulative and body-based practices are safe (n=234, 40.91%), effective (n=226, 39.51%), that they should be integrated into mainstream medicine (n=199, 34.79%), that insurance companies should cover the cost of this therapy (n=218, 38.18%), and that clinicians should receive training via formal education (n=185, 32.34%) (**Figure 7**). They also disagreed when asked if they would be comfortable counselling patients about whole medical systems (n=94, 30.92%), and if they would be comfortable recommending whole medical systems to their patients (n=101, 33.22%).

**Figure 7:**
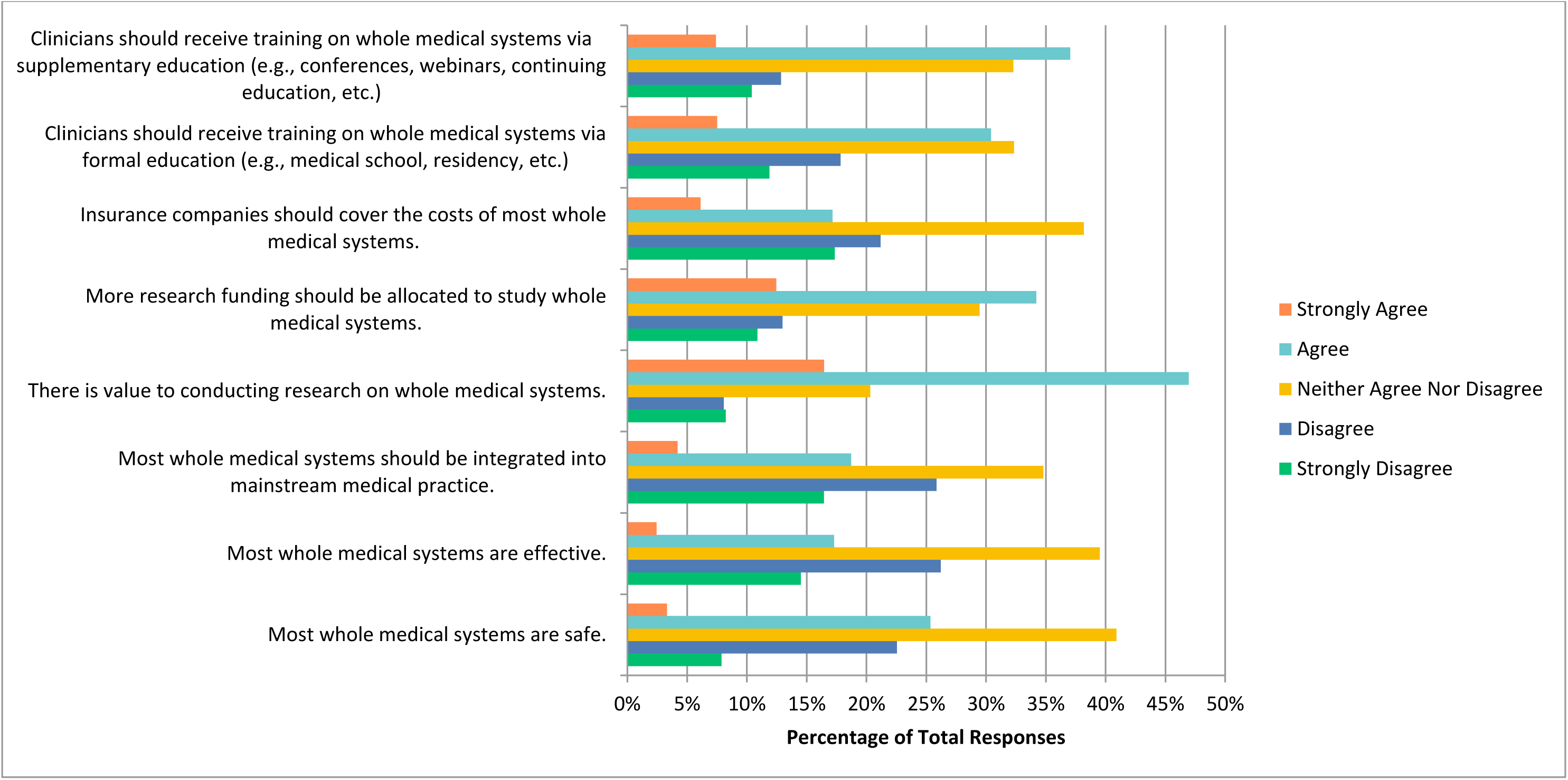
Agreement with the Following Statements Regarding Whole Medical Systems.

### Benefits and Challenges

The greatest benefits that respondents perceived to be associated with CAIM are ‘holistic approach to health and wellness’ (n=343, 60.07%), ‘empowerment of patients to take control of their own health’ (n=325, 56.02%), and ‘increased patient satisfaction and well-being’ (n=324, 56.74%) (**Figure 8**). On the other hand, the most challenging aspects respondents perceived to be associated with CAIM are ‘lack of scientific evidence for safety and efficacy’ (n=533, 92.70%), ‘lack of standardization in product quality and dosing’ (n=479, 83.30%), and difficulty in distinguishing legitimate practices from scams or fraudulent claims (n=423, 73. 57%) (**Figure 9**).

**Figure 8:**
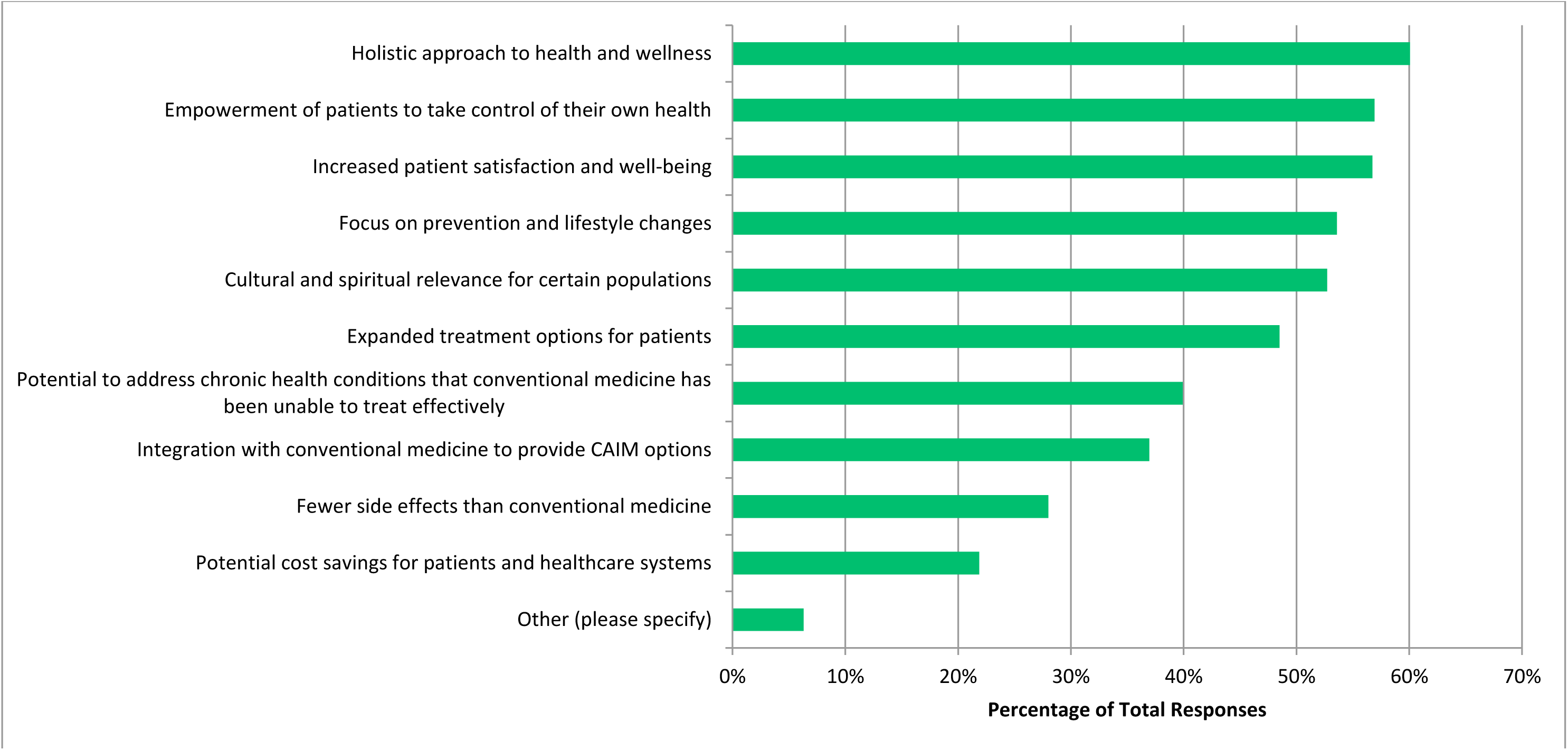
Benefits Perceived to be Associated with CAIM.

**Figure 9:**
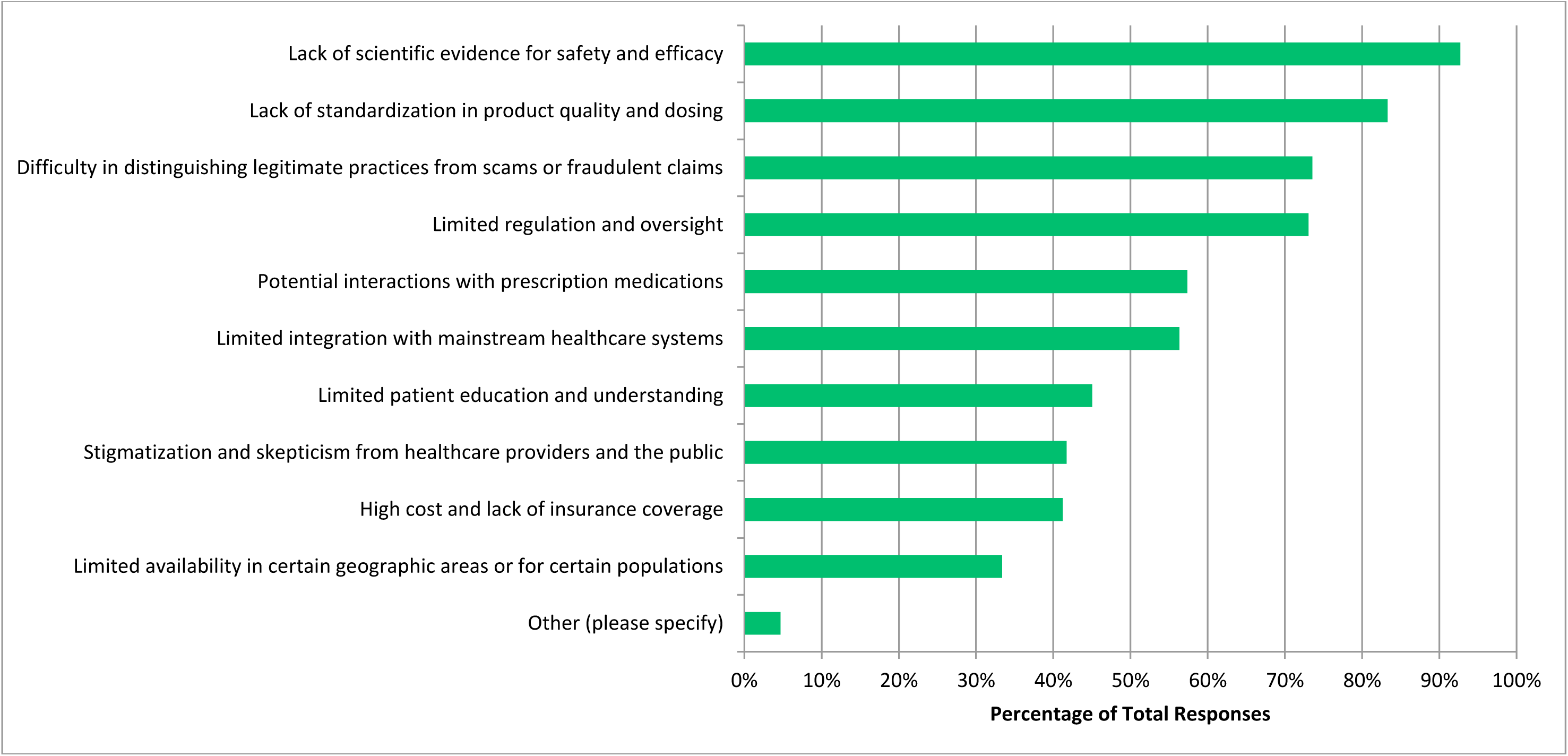
Challenges Perceived to be Associated with CAIM.

### Thematic Analysis

The results yielded 23 codes from the 115 open-ended responses that were received. From these codes, 5 distinct themes were created, which encompass the specific patterns that were established from the dataset. Firstly, “concerns regarding dangers associated with CAIM including false impressions and potential interactions” involved the potential negative consequences that respondents felt were associated with CAIM. Next, “disagrees and is not in support of CAIM” included responses that felt CAIM should not be used in oncology. Also, “more research is required” summarized responses who felt this area needed more evidence-based research to reach a consensus. “In support of specific CAIM for explicit purposes (e.g. emotional/supportive, not curative)” included responses who believed in the usefulness of CAIM for certain distinct purposes. Finally, “belief in integrative approaches in CAIM is beneficial” was indicative of those who believed a holistic approach involving both traditional and CAIM was ideal. Coding and thematic analysis data are available at: https://osf.io/4qny9.

## Discussion

The objective of this study was to collect the perceptions of oncology researchers and clinicians regarding CAIM. It was found that there are mixed perceptions regarding the various CAIM modalities, with more positive perceptions towards mind-body therapies and more negative feelings towards others such as biofield therapies. However, most respondents agreed that there is a need for more research in the field of CAIM in oncology.

### Comparative Literature

The results align with previous research that has explored healthcare processionals’ views on CAIM. In two similar studies conducted in a sample of psychiatry and neurology researchers and clinicians respectively, it was noted that most respondents perceived mind-body therapies to be the most promising CAIM, and while they agreed that most CAIM therapies in general are safe, many disagree that CAIM therapies are effective [48, 49]. A common theme noted in previous studies is that there is a lack of adequate evidence for the use of CAIM in oncological settings [50,51]. Additionally, previous studies have found that the risk of interactions between CAIM products and oncological treatment is a significant factor that they must consider in mitigating patient risk [52,53]. In our study, respondents also felt that these were major concerns for the use of CAIM in oncology, which may be the reason why they chose the option indicating that various CAIM modalities may not be safe and/or effective. Specifically, from the thematic analysis conducted, the theme with the most responses was “concerns regarding dangers associated with CAIM including false impressions and potential interactions” therefore demonstrating their concern. A previous study conducted which examined and compared the perceptions of traditional health care providers with Indigenous Australian healthcare providers in the field of oncology on the use of CAIM also found that the Indigenous providers had a greater understanding and openness towards CAIM [52]. Also, a study conducted in the field of paediatric oncology on opinions surrounding CAIM found that giving false hope to patients was a major concern for healthcare practitioners [54]. It is expected that due to the variability of different malignancies and their respective prognoses, clinicians would want to present their patients with the most evidence-based option for improvement. This is in line with the results of our study, as many respondents were unsure about the safety and effectiveness of some forms of CAIM.

Through this study, participants were asked about their perceptions on various CAIM therapies. It was noted that mind-body therapies and biologically based therapies received the most positive responses. Mind-body therapies such as yoga and meditation have been found to have positive outcomes on certain aspects of cancer diagnoses such as improving quality of life, reducing fatigue, improving sleep, and decreasing anxiety [55]. However, it is important to clarify that this modality is not associated with improving cancer outcomes; they merely improve associated symptoms. This could explain why respondents in our study were unsure when asked about the effectiveness of these therapies, and if they should be integrated into mainstream medical practices. Next, biologically based practices encompass modalities such as vitamins and supplements. Evidence-based guidelines are available for oncologists regarding which of these therapies are safe to be integrated into cancer care [56], which may be why the respondents in our study are more inclined to agree with this approach.

The respondents in this study consistently felt that there is value to conducting research in the field of CAIM, even across all modalities. Previous studies have also found that the lack of knowledge about CAIM can create a barrier to effective clinician-patient communication [57], due to fear of receiving less care from physicians if patients disclose their use of CAIM [54]. An additional study that compared the perceptions of physicians versus nurses on CAIM found that physicians had reservations regarding a lack of knowledge about the potential value of CAIM [58].

### Strengths and Limitations

One of the most impactful strengths of our study is that it was able to generalize the perceptions of oncological researchers and clinicians on CAIM, given the large and international sample of individuals who were surveyed. Moreover, during our data collection process, the names and email addresses of study participants were accessed based on the National Library of Medicine (NLM) categorization of such, thus ensuring that the email addresses of a wide variety of authors who have published articles in MEDLINE-indexed oncology journals had been collected. Our study also communicated multiple reminders to prospective study participants with the aim of improving the response rate. Moreover, our survey was sent to individuals who have published in oncology journals throughout the past approximately three years, thus limiting the potential for invalid/inactive email addresses.

Challenges and limitations faced by our study include the possibility of various biases. Nonresponse bias occurs when the characteristics of non-responders differ from responders and may thus have affected the sample of our study and the generalizability of the CAIM perspectives we collected [59]. Similarly, recall bias, which may arise due to individuals’ prior exposure to CAIM usage in oncology settings, may have also affected the generalizability of our findings [59]. Moreover, the survey within our study was written and administered in English. Therefore, it is more likely that English-speaking participants completed the survey, possibly having excluded or discouraged non-English clinicians and researchers from providing their perspectives on CAIM. The methodology of our study also presented various limitations. For example, due to our sampling strategy, which involved extracting the names and email addresses of authors of articles published in scholarly oncology journals, it is likely that our study sampled proportionally more researchers than clinicians. In addition, the response rate reported within our study was likely an underestimation of the true value of such. This limitation is enforced by the potential for inactive and invalid email addresses, which may have been a result of changing professions/employers, retiring, or passing away, in addition to the potential that the individuals may have been unavailable during the defined study period due to reasons such as vacations/leaves of absence. Finally, another constraint to bear in mind is that CAIM is a broad term. Despite our classification into five categories (mind-body therapies, biologically based therapies, manipulative and body-based practices, biofield therapies, and whole medical systems), the safety and efficacy profiles vary for each therapy [16]. Consequently, participants were required to formulate generalized opinions on these therapies rather than offering specific insights for each distinct type.

## Conclusions

The aim of this study was to understand the perspectives of oncology researchers and clinicians regarding various CAIM therapies. Participants were given the opportunity to rank their views on numerous therapies and offer additional insights they deemed relevant. This yielded valuable understanding into the current perceptions of CAIM within the field of oncology, serving as a foundation for future research, as indicated by the inclination of respondents. These findings also demonstrate potential for the development of customized educational resources, given respondent agreement on the potential benefits of further education. While past literature has identified patient interest in CAIM, this study stands as the first to specifically address this research question and provide insights into the perceptions of oncology researchers and clinicians. It is anticipated that the outcomes and analysis presented in this study can make meaningful contributions to both the fields of oncology and CAIM, establishing a groundwork for subsequent research endeavours.

## List of Abbreviations

CAIM: complementary, alternative, and integrative medicine

NCCIH: National Centre for Complementary and Integrative Health

NLM: National Library of Medicine

PMIDs: PubMed Identifiers

## Declarations

### Ethics Approval and Consent to Participate

This study received approval from the University Tübingen Research Ethics board before commencement (REB Number: 389/2023BO2).

## Consent for Publication

All authors consent to this manuscript’s publication.

## Availability of Data and Materials

All data and materials associated with this study have been posted on the Open Science Framework and can be found here: https://doi.org/10.17605/OSF.IO/FV62Y

## Authors’ Contributions

JYN: designed and conceptualized the study, collected and analysed data, drafted the manuscript, and gave final approval of the version to be published.

JK: assisted with the collection and analysis of data, made critical revisions to the manuscript, and gave final approval of the version to be published.

HC: assisted with the design and concept of the study and the analysis of data, made critical revisions to the manuscript, and gave final approval of the version to be published.

## Data Availability

All data and materials associated with this study have been posted on the Open Science Framework.

https://doi.org/10.17605/OSF.IO/FV62Y

## Acknowledgements

We gratefully acknowledge Nadia Dorca for her contributions to the protocol and data collection.

